# AVIFAVIR for Treatment of Patients with Moderate COVID-19: Interim Results of a Phase II/III Multicenter Randomized Clinical Trial

**DOI:** 10.1101/2020.07.26.20154724

**Authors:** Andrey A. Ivashchenko, Kirill A. Dmitriev, Natalia V. Vostokova, Valeria N. Azarova, Andrew A. Blinow, Alina N. Egorova, Ivan G. Gordeev, Alexey P. Ilin, Ruben N. Karapetian, Dmitry V. Kravchenko, Nikita V. Lomakin, Elena A. Merkulova, Natalia A. Papazova, Elena P. Pavlikova, Nikolay P. Savchuk, Elena N. Simakina, Tagir A. Sitdekov, Elena A. Smolyarchuk, Elena G. Tikhomolova, Elena V. Yakubova, Alexandre V. Ivachtchenko

## Abstract

In May 2020 the Russian Ministry of Health granted fast-track marketing authorization to RNA polymerase inhibitor AVIFAVIR (favipiravir) for the treatment of COVID-19 patients. In the pilot stage of Phase II/III clinical trial, AVIFAVIR enabled SARS-CoV-2 viral clearance in 62.5% of patients within 4 days, and was safe and well-tolerated.

## INTRODUCTION

The pandemic of the novel coronavirus infection (COVID-19) represents an unprecedented disaster for healthcare providers and economy worldwide. The urgent requirement for the effective treatments has sparked an intense effort on the part of the pharmaceutical industry and shifted drug development to a new scale of commitment and collaboration.

Repositioning of antiviral drugs that could be effective against SARS-CoV-2 is one of the common strategies in the fight against COVID-19. In May 2020, remdesivir received conditional approval from U.S. Food and Drug Administration for the treatment of hospitalized patients with severe COVID-19. However, there are still limited data on its efficacy and safety and limitations of the intravenous route of administration [1]. Thus, the development of the effective oral antiviral agents for use at earlier stages of the disease is still warranted.

Favipiravir is an RNA-dependent RNA polymerase inhibitor marketed in Japan (Avigan) and China (Favilavir) as a second-line treatment of novel or re-emerging influenza outbreaks [2]. Earlier this year it was reported to demonstrate antiviral activity against SARS-CoV-2 in Vero E6 cells (EC_50_, 61.88 μM, CC_50_ > 400 μM, SI > 6.46) and to provide shorter viral clearance time in patients with COVID-19 [3]. A non-randomized study conducted in China demonstrated a median viral clearance time of 4 days, versus a period of 11 days for lopinavir/ritonavir (p < 0.001) [4]. The World Health Organization (WHO) also listed Favipiravir as a candidate experimental treatment (broad spectrum antiviral) [5].

Favipiravir under brand name AVIFAVIR was resynthesized and developed in Russia by the joint venture of Russian Direct Investment Fund and ChemRar Group. Following preclinical tests, a Phase II/III clinical trial of AVIFAVIR in patients with COVID-19 was initiated. This brief report provides the interim results of the study from 60 patients enrolled in the pilot stage (Phase II).

## METHODS

This was an adaptive, multicenter, open label, randomized, Phase II/III clinical trial of AVIFAVIR versus standard of care (SOC) in hospitalized patients with moderate COVID-19 pneumonia. The study design was based on WHO R&D Blueprint recommendations [6]. The pilot stage of the study was conducted in April and May 2020 at six Russian clinical sites in Moscow, Smolensk, and Nizhniy Novgorod (ClinicalTrials.gov Identifier: NCT04434248). The Independent Data Monitoring Committee (IDMC) was introduced to review the results of the interim analysis.

The eligible patients included hospitalized men and non-pregnant women of 18 years or older who signed the informed consent form, had moderate PCR-confirmed COVID-19 (positive test at screening), were able to administrate the drug orally and willing to use adequate contraception during the study and 3 months after its completion.

The patients were randomized at a 1:1:1 ratio to receive either AVIFAVIR 1600 mg BID on Day 1 followed by 600 mg BID on Days 2-14 (1600/600 mg), or AVIFAVIR 1800 mg BID on Day 1 followed by 800 mg BID on Days 2-14 (1800/800 mg), or SOC according to the Russian guidelines for treatment of COVID-19 [7]. During the study, patients in all groups were allowed to use pathogenetic and symptomatic treatment; patients in the AVIFAVIR groups were not allowed to use other antivirals or antimalarial drugs.

The objective of the pilot stage of the study was to preliminary assess the efficacy and safety of AVIFAVIR, and to select the optimal dosing regimen for further evaluation at the pivotal stage (Phase III). The primary efficacy endpoint at the pilot stage was the elimination of SARS-CoV-2 by Day 10 (defined as two negative PCR tests with at least a 24-hour interval). Qualitative Real-Time RT-PCR tests were performed at local laboratories of the clinical sites or in the central laboratory of Rospotrebnadzor.

The study assessments included daily vital signs, SpO_2_ and WHO Ordinal Scale for Clinical Improvement (WHO-OSCI); PCR for SARS-CoV-2 detection in nasopharyngeal and/or oropharyngeal swabs at baseline and on Days 5, 10, and 15; chest computed tomography (CT) scan at baseline and on Day 15; physical examination, complete blood count, biochemistry, C-reactive protein (CRP), urinalysis, and electrocardiogram at baseline and on Days 5 and 15. If not discharged from the hospital earlier, the patients attended follow-up visits on Day 22 and Day 29. All patients were followed until Day 29.

The “go-no-go” decision to start the pivotal stage of the study was based on the exact single-stage Phase II assessment at one-sided α=0.05 and 80% power [8]. 13/18 (72.2%) or more patients would have been sufficient to demonstrate that the viral clearance in 80% of patients by Day 10 was plausible, and that the response was greater than the presumably non-effective level of 50%. The secondary endpoints that were assessed during the interim analysis were the rate of viral clearance by Day 5, time to normalization of clinical symptoms (i.e. body temperature), changes on CT scan by Day 15, and incidence and severity of adverse events related to the study drug.

## RESULTS

Upon signing the informed consent form and confirmed eligibility criteria, 60 patients hospitalized with COVID-19 pneumonia were randomized into three treatment groups: AVIFAVIR 1600/600 mg, AVIFAVIR 1800/800 mg, or SOC. Each group comprised 20 patients and all randomized patients constituted safety and intent-to-treat (ITT) analysis sets.

The AVIFAVIR and control groups were generally comparable in demographic and baseline characteristics. 28/60 (46.7%) had risk factors for severe disease (i.e. age 60 and older and/or concurrent chronic conditions); 45/60 (75.0%) were on ambient air (Score 3 on WHO-OSCI) and 15/60 (25.0%) required supplemental oxygen via mask or nasal cannula (Score 4 on WHO-OSCI); 15/60 (25.0%) had body temperature > 38°C, and 42/60 (70.0%) had CRP > 10 mg/L. Mean disease duration at baseline was 6.7 days from the start of the symptoms.

In the AVIFAVIR groups, the study drug was administered for a mean period of 10.9 ± 2.8 days. In the SOC group, hydroxychloroquine or chloroquine was administered to 15/20 (75.0%) patients, lopinavir/ritonavir was used in 1/20 (5%) patient, and 4/20 (20%) patients did not receive etiotropic treatment. The concomitant therapy of COVID-19 in all groups included antibiotics, anticoagulants and/or immunosuppressants, as well as symptomatic treatment.

Both dosing regimens of AVIFAVIR demonstrated similar virologic response. On Day 5, the viral clearance was achieved in 25/40 (62.5%) patients on AVIFAVIR and in 6/20 (30.0%) patients on SOC (p=0.018). By Day 10 the viral clearance was achieved in 37/40 (92.5%) patients on AVIFAVIR and in 16/20 (80.0%) patients on SOC (p=0.155). Thus, the required number of responders to demonstrate proof of concept was attained.

The median time to body temperature normalization (< 37°C) was 2 days (IQR 1-3) in the AVIFAVIR groups and 4 days (IQR 1-8) in the SOC group (p=0.007). By Day 15, chest CT scans improved in 36/40 (90.0%) patients on AVIFAVIR and 16/20 (80.0%) patients on SOC (p=0.283).

Adverse drug reactions to AVIFAVIR were reported in 7/40 (17.5%) patients, including diarrhea, nausea, vomiting, chest pain and an increase in liver transaminase levels. The adverse drug reactions were mild to moderate and caused early discontinuation of the study drug in 2/40 (5.0%) patients.

Two patients on AVIFAVIR 1600/600 mg were moved to intensive care unit, received mechanical ventilation and later died. Both patients had the increased risk of severe disease, including diabetes mellitus, arterial hypertension, obesity, CRP > 50 mg/L, and supplemental oxygen at baseline. 13/20 (65.0%) patients on AVIFAVIR 1600/600 mg, 17/20 (85.0%) patients on AVIFAVIR 1800/800 mg and 17/20 (85.0%) patients on SOC were discharged from the hospital and/or achieved Score 2 on WHO-OSCI by Day 15.

## DISCUSSION

AVIFAVIR demonstrated rapid antiviral response against SARS-CoV-2. The proportion of patients who achieved negative PCR on Day 5 on both dosing regimens of AVIFAVIR was twice as high as in the control group (p<0.05). The median loading dose of AVIFAVIR given to responders was 43.9 mg/kg (IQR 40.0-47.1), and to those with positive PCR on Day 5, it was 39.1 mg/kg (IQR 35.6-43.9). There were no new safety concerns related to AVIFAVIR as all adverse reactions were mild to moderate in severity and were consistent with those reported previously for AVIGAN [2, 9]. No increasing toxicity was observed in patients who received higher doses of AVIFAVIR. Based on this observation, it was decided to introduce a weight-based dosing regimen of AVIFAVIR in the pivotal stage of the study with the target loading dose of ≥ 44 mg/kg and the treatment duration of up to 10 days.

While clinical effects of AVIFAVIR will be further studied in the pivotal stage of the study, the pilot data demonstrates proof of concept and high treatment potential of the drug in moderately ill patients. It also suggests that the administration of AVIFAVIR to patients with signs of respiratory distress and cytokine storm should be considered only as part of the combination therapy that includes anti-inflammatory agents with proven efficacy against COVID-19.

Based on the interim results of the Phase II/III clinical trial, the Russian Ministry of Health granted a conditional marketing authorization to AVIFAVIR, which makes it the only approved oral drug for treatment of moderate COVID-19 to date.

## Data Availability

N/A

## Acknowledgements

The authors thank the co-investigators MD Oksana Y. Shaydyuk of City Clinical Hospital n.a. O.M. Filatov (Moscow, Russia); MD, PhD Dmitry A. Garkavi and MD Anna V. Bushmanova of First Moscow State Medical University n.a. I.M. Sechenov (Moscow, Russia); MD, PhD Anton V. Potapenko and MD Anna G. Sorokina of Moscow State University n.a. M. V. Lomonosov (Moscow, Russia); MD PhD Olga S. Rozinkova of Clinical hospital No.1 (Smolensk, Russia); MD, PhD Veronika S. Vasilieva, MD Ekaterina E. Shokhina, MD Natalia O. Simonova, and MD Valery A. Luybinskiy of Central Clinical Hospital with Polyclinic (Moscow, Russia), as well as members of the Independent Data Monitoring Committee, including MD, PhD, Professor Alexey V. Kravchenko of Central Research Institute of Epidemiology (Moscow, Russia), MD, PhD, Professor, Russian Academy of Sciences Professor Kirill A. Zykov of Moscow State Medical Dental University n.a. A.I. Yevdokimov (Moscow, Russia) and MD, PhD, Professor Vladimir V. Rafalskiy of Immanuel Kant Baltic Federal University (Kaliningrad, Russia).

## Authors’ contributions

EPP, NVL, EAS, IGG, ENS, EGT were Principal Investigators responsible for recruitment of patients, study treatment, and data collection in compliance with the Protocol and ICH GCP. AAI, KAD and TAS conceived this project, proposed a variant of its organization and controlled the progress of its implementation. NVV, VNA and ANE developed the clinical trial protocol, worked on the statistical aspects of the study and the analysis of the results. EAM, AAB and EVY organized the clinical trial and collection of the study data. RNK developed the preclinical study design and organized its implementation. API, DVK developed the technology and organized the production of the substance. NAP developed the composition and technology for the production of AVIFAVIR tablets. NPS studied and analyzed the possible market for AVIFAVIR. AVI carried out scientific management of the project and edited the publication.

---

**Figure 1.**
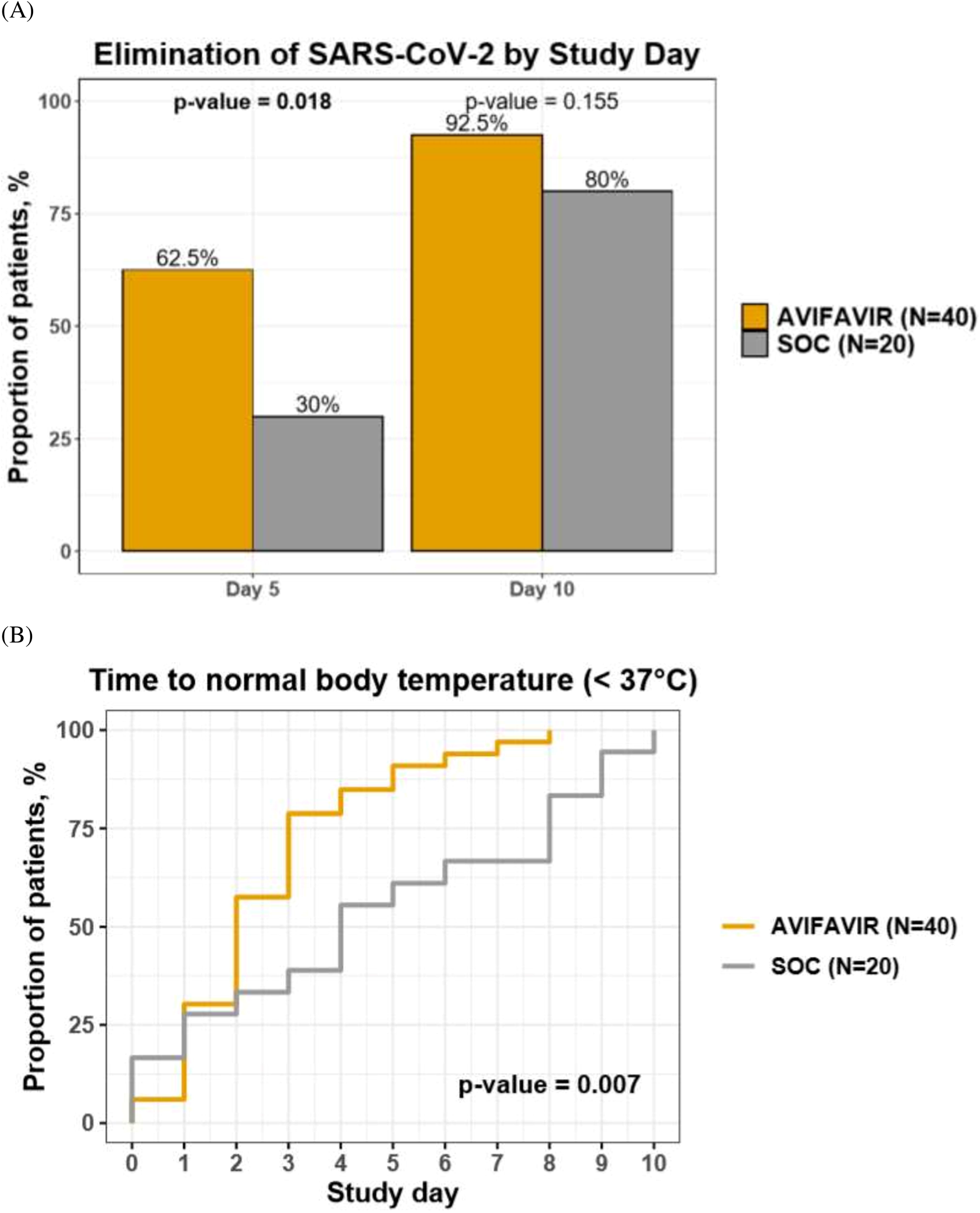
*A*. Elimination of SARS-CoV-2 in nasopharyngeal and/or oropharyngeal swabs of patients treated with AVIFAVIR or SOC on Day 5 and Day 10 of the study treatment. *B*. Time to normal body temperature in patients treated with AVIFAVIR or SOC (Kaplan-Meier curves).

